# Causes of death in people with cardiovascular disease: a prospective UK Biobank cohort study

**DOI:** 10.1101/2021.03.26.21254418

**Authors:** Michael Drozd, Mar Pujades-Rodriguez, Fei Sun, Kevin N Franks, Patrick J Lillie, Klaus K Witte, Mark T Kearney, Richard M Cubbon

## Abstract

Cardiovascular disease (CVD) mortality has substantially improved over recent decades. Some evidence indicates this has been paralleled by an increasing proportion of non-cardiovascular mortality in people with CVD. However, the contemporary causes of death across a broad spectrum of CVDs, either alone or in combination, remains unclear.

We analysed cardiovascular, infection, cancer and other causes of death prior to the COVID-19 pandemic in 493,280 participants in the prospective UK Biobank study. Studied CVDs included baseline: abdominal aortic aneurysm, atrial fibrillation, coronary artery disease, heart failure, hypertension, peripheral vascular disease, stroke, valvular heart disease and venous thromboembolic disease; we separately considered cardiovascular multimorbidity defined as the total number of these baseline CVDs. Crude mortality rates and Poisson regression analysis were used to quantify the absolute and relative risk of cause-specific death. Associations are reported as incidence rate ratios (IRR) with 95% CIs.

During a median follow-up of 10.9 [IQR 10.1-11.6] years per participant, there were 27,729 deaths (20.4% primarily attributed to CVD, 53.6% to cancer, 5.0% to infection and 21.0% to other causes). As the number of co-morbid CVDs increased, the proportion of cardiovascular and infection-related deaths increased, whereas cancer and other deaths decreased. Accrual of multiple CVDs was associated with marked increases in relative risk of infection and cardiovascular death; versus those without CVD, people with three or more CVDs, the relative risk of cardiovascular death increased most (IRR 3.89; 95%CI 3.59-4.21), followed by infection (4.41; 3.44-5.64), with other (2.01; 1.72-2.35) and cancer (1.52; 1.35-1.72) being substantially less increased. All studied CVDs except atrial fibrillation were independently associated with increased risk of infection death, with heart failure (2.73; 1.60-4.66) and valvular heart disease (3.09; 2.38-4.00) posing the greatest risk.

In conclusion, causes of death vary substantially between differing baseline CVDs, and according to the number of baseline CVDs, with non-cardiovascular deaths due to cancer and infection making an important contribution. Holistic and personalized care are likely to be important tools for continuing to improve outcomes in people with CVD.

## Main

Cardiovascular disease (CVD) mortality has substantially improved over recent decades.^1^ Some evidence indicates this has been paralleled by an increasing proportion of non-cardiovascular mortality in people with CVD; for example, non-cardiovascular death now accounts for approximately 40% of deaths in people with chronic heart failure with reduced left ventricular ejection fraction.^2^ However, the contemporary causes of death across a broad spectrum of CVDs, either alone or in combination, remains unclear, hindering the planning of strategies that continue improving outcomes in people with CVD.

We addressed this using the prospective UK Biobank study, which recruited 502,505 United Kingdom residents aged 37–73 years between 2006-10. Detailed methods for our analysis are described in the appendix (pp 8-10). Briefly, we excluded 9,225 (1.8%) participants due to missing baseline data, loss to follow-up, or withdrawal of consent (appendix pp 8-10). We extracted the primary cause of death, coded according to ICD10 from death certification data and classified these as cardiovascular, cancer, infection or other (appendix pp 9). Crude mortality rates and Poisson regression analysis were used to quantify the absolute and relative risk of death. Relative risks of death, presented as incidence rate ratio (IRR) with 95% confidence intervals, are adjusted for age, sex, ethnicity, socio-economic deprivation, smoking status, obesity, and self-reported: diabetes, cancer, respiratory, liver, kidney, neurological, psychiatric and rheumatological disease.

Among 493,280 participants, 131,202 (26.6%) had one self-reported CVD (within the nine studied CVDs), 21,605 (4.4%) had two CVDs and 3,561 (0.7%) had three of more CVDs. Within these, there were 130,792 (26.5%) with hypertension, 22,847 (4.6%) with coronary artery disease, 12,386 (2.5%) with venous thromboembolic disease, 6,996 (1.4%) with stroke, 4,600 (0.9%) with valvular disease, 3,649 (0.7%) with atrial fibrillation/flutter, 3,160 (0.6%) with peripheral vascular disease, 781 (0.2%) with heart failure and 418 (0.1%) with abdominal aortic aneurysm.

During a median follow-up period of 10.9 [IQR 10.1-11.6] years per participant, there were 27,729 deaths (censored 31/12/2019, prior to the COVID-19 pandemic); of these, 5,648 (20.4%) were primarily attributed to CVD, 14,684 (53.6%) to cancer, 1,385 (5.0%) to infection and 5,832 (21.0%) to other causes.

In participants with one CVD, only 22.4% of deaths were attributed to CVD, whereas 50.5% were attributed to cancer, 5.7% to infection, and 21.4% to other causes (**Figure 1A**). As the number of co-morbid CVDs increased, the proportion of cardiovascular and infection-related deaths increased, whereas cancer and other deaths decreased. Indeed, in people with three or more CVDs, 43.1% of deaths were attributed to CVD; absolute mortality rates are presented in appendix pp 11. Since the characteristics of people with increasing numbers of comorbid CVDs will differ, we also examined the adjusted risk of cardiovascular, cancer, infection or other death, relative to people without baseline CVD (Figure 1A and appendix pp 12). As expected, the presence of one baseline CVD modestly increased the risk of cardiovascular death, with small increases in the risk of cancer or other death. Surprisingly, infection death was increased to a similar extent to cardiovascular death. In people with three or more CVDs (versus no CVD), the relative risk of cardiovascular death increased most (IRR 7.00; 6.24-7.84), followed by infection (4.41; 3.44-5.64), with other (2.01; 1.72-2.35) and cancer (1.52; 1.35-1.72) being substantially less increased. Hence, accruing baseline cardiovascular comorbidity is particularly associated with increasing relative and absolute importance of both cardiovascular and infection death.

**Figure 1:**
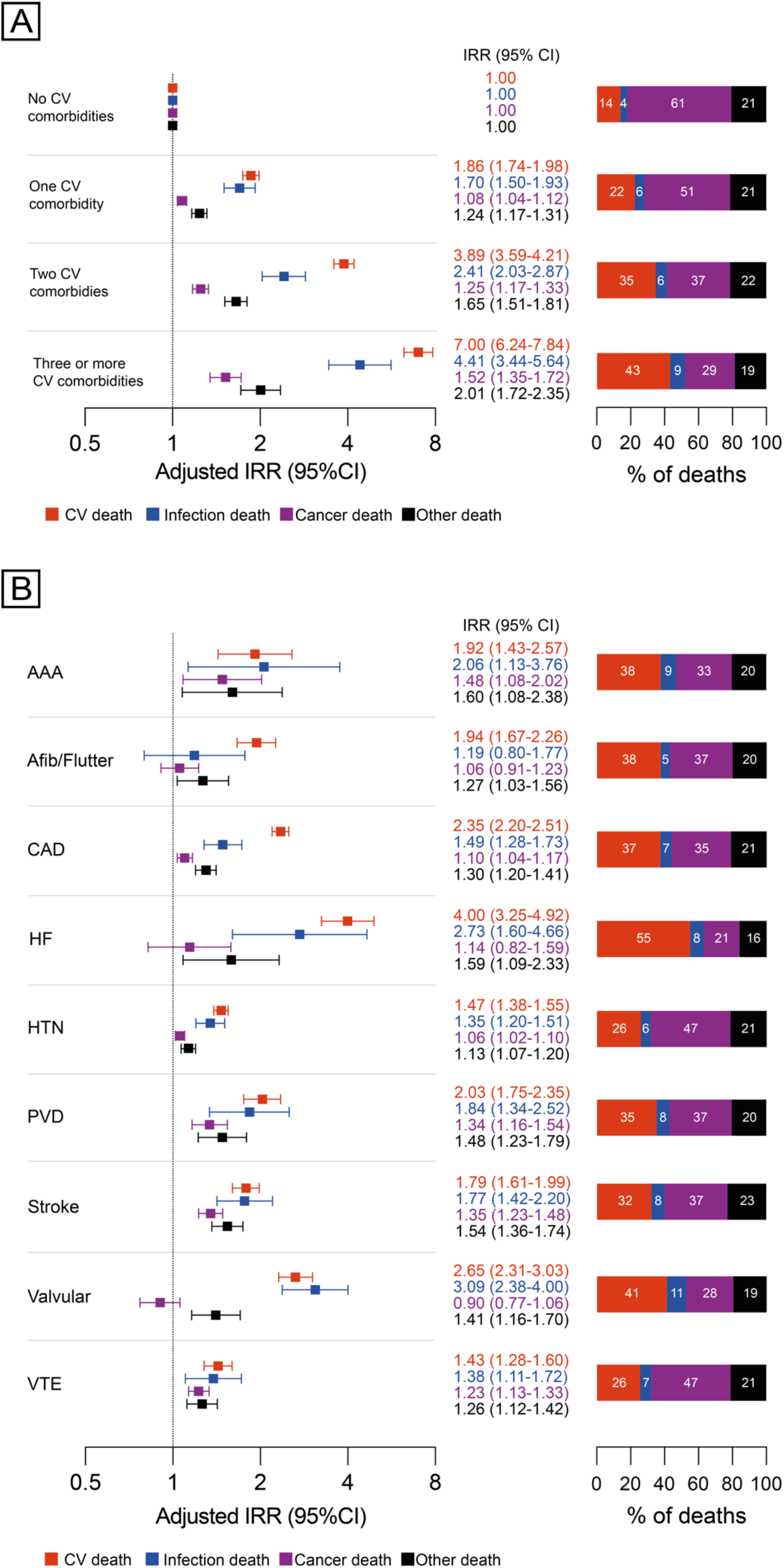
Causes of death according to baseline CVDs. Forest plots illustrate adjusted IRRs and 95% confidence intervals (plotted on a log2 scale) for cardiovascular, infection, cancer and other death according to number of baseline CVDs (**A**) or specific baseline CVDs (**B**) using multivariate Poisson regression analysis; bar charts illustrate the absolute percentage of deaths attributed to each cause. AAA: abdominal aortic aneurysm; Afib: atrial fibrillation; flutter: atrial flutter; CAD: coronary artery disease; HF: heart failure; HTN: hypertension; PVD: peripheral vascular disease; VTE: venous thromboembolism; IRR: incidence rate ratio; CI: confidence interval

Next, we explored how specific baseline CVDs were associated with cause of death (**Figure 1B** and appendix pp 13) and found substantial variation in the absolute proportion of cardiovascular death, and the relative risk of cardiovascular death (versus people without that disease), across CVD types. For example, cardiovascular death predominated in people with heart failure, whereas cancer death was most common in people with hypertension and venous thromboembolic disease. Similarly, the relative risk of cardiovascular death was much larger in people with heart failure (4.00; 3.25-4.92), than hypertension (1.47; 1.38-1.55) or venous thromboembolic disease (1.43; 1.28-1.60). Despite the wider estimated confidence intervals in this analysis, it is apparent that the relative risk of infection death was also substantially elevated in people with heart failure (2.73; 1.60-4.66) and valvular heart disease (3.09; 2.38-4.00), and was elevated in people with all baseline CVDs except atrial fibrillation/flutter

These data have important clinical implications. Firstly, cancer is a common cause of death in people with CVD (cancer types presented in appendix pp 14), and lifestyle interventions for CVD might also reduce cancer risk so this should be emphasised to patients.^3^ Secondly, the importance of cardio-oncology is emphasised by the common occurrence of cancer death in people with CVD. Thirdly, the increasing absolute and relative risk of infection death in people with increasing cardiovascular multimorbidity suggests we need to better understand and address mechanisms of infection risk in people with CVD.^4^ In conclusion, holistic and personalized care are likely to be important tools for continuing to improve outcomes in people with CVD.

## Data Availability

The UK Biobank dataset is available to all bona fide researchers for all types of health-related research which is in the public interest

## Acknowledgments

This research was conducted using the UK Biobank resource, under approval 59585. MD is supported by a British Heart Foundation Clinical Research Training Fellowship (FS/18/44/33792). RMC was supported by a British Heart Foundation Intermediate Clinical Research Fellowship (FS/12/80/29821). MTK is a British Heart Foundation Professor. MPR is currently employed by IQVIA, a contract research organisation. At no time did any authors nor their institutions receive other payment or services from a third party for any aspect of the submitted work.

## Appendix

### Methods

#### Study design and data collection

The UK Biobank study is a prospective observational study that recruited 502,505 residents in the United Kingdom aged 37-73 years between 2006-2010. This resource was developed with funding provided by the UK Government and biomedical research charities with the aim to improve understanding of disease. All researchers are able to apply for access to this resource. Detailed information for study design and conduct are available at the UK Biobank website (https://www.ukbiobank.ac.uk). Participants attended one of 22 assessment centres across the United Kingdom. Biological measurements were recorded at baseline, and participants completed a touchscreen and nurse-led interview, as previously described.^1^ UK Biobank received ethical approval from the NHS Research Ethics Service and this analysis was undertaken under application number 59585. Participants provided written informed consent and are free to withdraw from UK Biobank at any time and request that their data no longer be used.

#### Assessment of demographic factors and morbidity

At recruitment age, sex, ethnicity and socioeconomic status were collected at study recruitment by UK Biobank. Ethnicity was participant-classified within pre-defined categories by UK Biobank including White, Mixed, Asian or British Asian, Black or British Black, Chinese, or other ethnic groups. Self-reported smoking status was recorded at recruitment defined as never, former or current. We categorised the Townsend score collected at recruitment into quintiles. Obesity was classified according to body-mass index recorded at baseline based on the WHO’s definitions: class 1 (30.0–34.9 kg/m^2^), class 2 (35.0–39.9 kg/m^2^), and class 3 (≥ 40 kg/m^2^). Medical conditions and operations were self-reported at study recruitment during a face-to-face interview with a nurse. We classified these into disease groups (pp 16). In addition to a broad range of cardiovascular comorbidities (abdominal aortic aneurysm, atrial fibrillation/flutter, coronary artery disease, heart failure, hypertension, peripheral vascular disease, stroke, valvular disease, venous thromboembolic disease), we also classified a broad range of other co-morbidities based on our previously published work to incorporate the information into our models (respiratory disease, diabetes, cancer (previous or current), chronic liver disease, chronic kidney disease, other neurological disease (not stroke), psychiatric disorder and chronic inflammatory and autoimmune rheumatic disease)^2^. The number of cardiovascular comorbidities (amongst those listed) was also calculated for each participant.

We excluded a total of 9,225 (1.8%) participants because of missing baseline data or long-term follow-up data, or withdrawal of consent from study. These included exclusions due to missing data for comorbidities (863 participants), BMI (3106 participants), smoking (2949), ethnicity (2777), socioeconomic deprivation (624 participants) and individuals lost to follow-up or who withdrew consent (1314); some participants had more than one variable missing.

#### Mortality ascertainment

The UK Biobank data portal provided mortality information for participants. This resource utilises linked national death registry data from NHS digital for participants in England and Wales, and from the NHS central register for participants in Scotland. In our analysis, we censored deaths to the 31^st^ December 2019 to ensure this was before the first recorded case of COVID19 in the UK.^3^ We extracted the underlying (primary) cause of death, coded according to the International Classification of Diseases (tenth revision) from death certification data; classified as cardiovascular (I00-I99, excluding infection codes: [I32.0, I32.1, I33.0, I33.9, I38, I39, I40.0, I41.0-I41.2, I43.0, I52.0-I52.1, I68.1, I98.1]), cancer (C00-C97), infection (as we have previously defined^2^) and other causes (any death that is not defined as cardiovascular, infection or cancer). We also conducted sensitivity analyses with alternative classification of cardiovascular death as I00-I99 (without any exclusions) and infection codes (excluding any code beginning with the letter I) revealing comparable estimates (pp 15).

#### Statistical analysis

Categorical variables are presented as number (%). Adjusted cause-specific mortality incidence rate ratios (IRRs) were estimated using Poisson regression models with exposure time modelled. Models were adjusted for all covariates including: age, sex, socioeconomic deprivation (based on IMD quintile), smoking status, obesity, respiratory disease, diabetes, cancer (previous or current), liver disease, kidney disease, other neurological disease, psychiatric disorder, rheumatological disease, abdominal aortic aneurysm, atrial fibrillation/flutter, coronary artery disease, heart failure, hypertension, peripheral vascular disease, stroke, valvular disease and venous thromboembolic disease. To assess the association of cardiovascular multimorbidity with cause of death, irrespective of the specific cardiovascular morbidities (abdominal aortic aneurysm, atrial fibrillation/flutter, coronary artery disease, heart failure, hypertension, peripheral vascular disease, stroke, valvular disease, venous thromboembolic disease), we modelled the number of cardiovascular comorbidities groups into four groups: none, one, two, three or more CV conditions. Age was modelled using restricted cubic splines with four knots for cardiovascular death, cancer death and other death, and five knots for infection death analyses, because these provided the best fit as assessed by the Akaike information and the Bayesian criterion (models including categorical, linear, or restricted cubic splines with three, four, and five knots and first-degree and second-degree fractional polynomials were compared). All tests were two-sided and statistical significance was defined as p<0·05. All statistical analyses were done with Stata/MP (version 16.1).

**Table 1:**
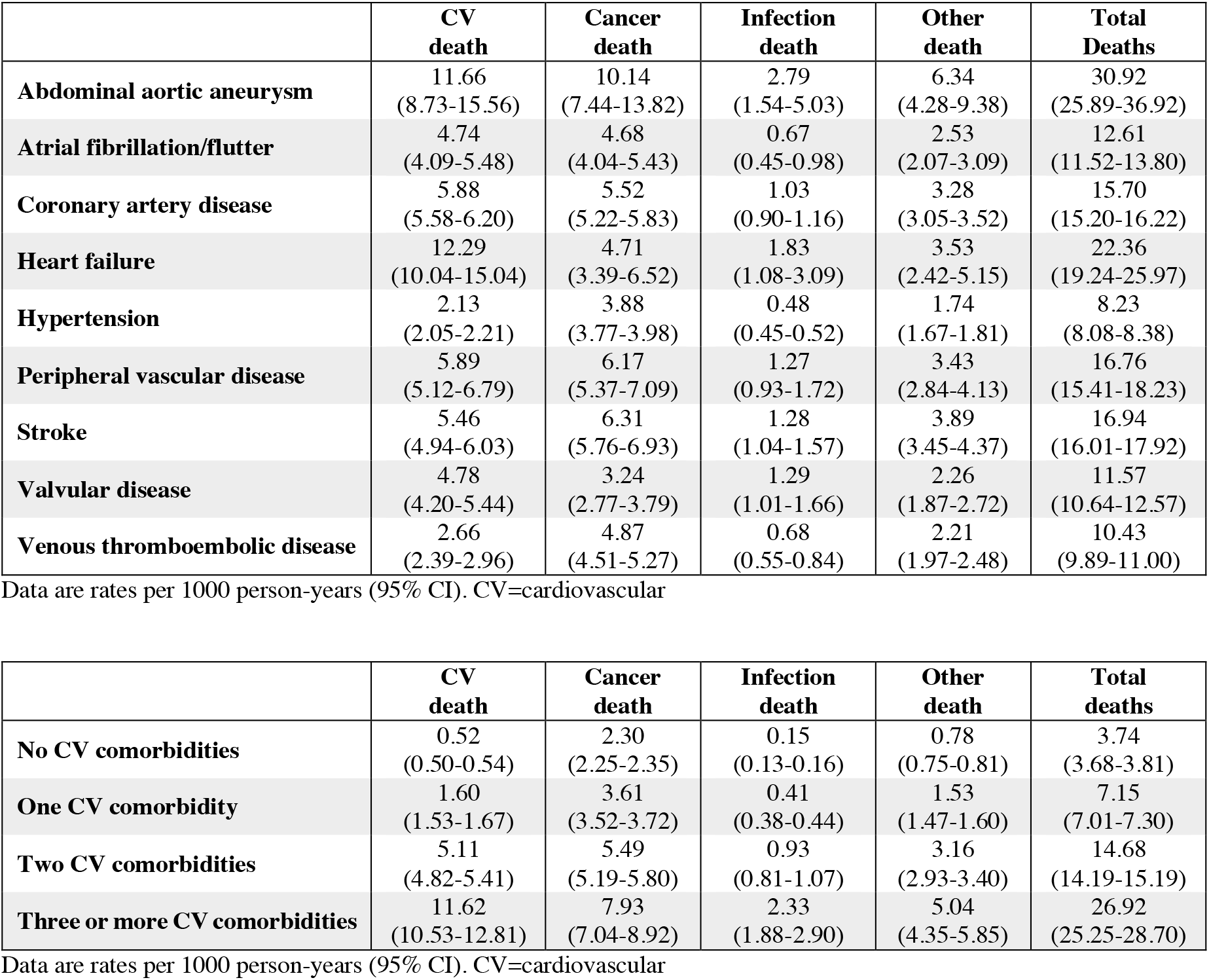
**Crude mortality rates per 1000 person-years of follow-up according to baseline cardiovascular comorbidities**

**Table 2:** **Causes of death according to number of baseline cardiovascular comorbidities**

|  | IRR (95% CI) |  |  |  |
| --- | --- | --- | --- | --- |
|  | CV death | Cancer death | Infection death | Other death |
| **CV comorbidities (reference: none)** |  |  |  |  |
| One CV comorbidity | 1.86<br>(1.74-1.98) | 1.08<br>(1.04-1.12) | 1.70<br>(1.50-1.93) | 1.24<br>(1.17-1.31) |
| Two CV comorbidities | 3.89<br>(3.59-4.21) | 1.25<br>(1.17-1.33) | 2.41<br>(2.03-2.87) | 1.65<br>(1.51-1.81) |
| Three or more CV comorbidities | 7.00<br>(6.24-7.84) | 1.52<br>(1.35-1.72) | 4.41<br>(3.44-5.64) | 2.01<br>(1.72-2.35) |

\*Fully adjusted models include age (modelled by use of restricted cubic splines with five knots for infection death, four knots for all other analyses), sex, socioeconomic deprivation, ethnicity, smoking, obesity, respiratory disease, diabetes, cancer, liver disease, kidney disease, neurological disease, psychiatric disease, rheumatological disease. CV=cardiovascular

**Table 3:** **Causes of death according to baseline cardiovascular comorbidities**

|  | IRR (95% CI) |  |  |  |
| --- | --- | --- | --- | --- |
|  | CV death | Cancer death | Infection death | Other death |
| **Abdominal aortic aneurysm** | 1.92<br>(1.43-2.57) | 1.48<br>(1.08-2.02) | 2.06<br>(1.13-3.76) | 1.60<br>(1.08-2.38) |
| **Atrial fibrillation/flutter** | 1.94<br>(1.67-2.26) | 1.06<br>(0.91-1.23) | 1.19<br>(0.80-1.77) | 1.27<br>(1.03-1.56) |
| **Coronary artery disease** | 2.35<br>(2.20-2.51) | 1.10<br>(1.04-1.17) | 1.49<br>(1.28-1.73) | 1.30<br>(1.20-1.41) |
| **Heart failure** | 4.00<br>(3.25-4.92) | 1.14<br>(0.82-1.59) | 2.73<br>(1.60-4.66) | 1.59<br>(1.09-2.32) |
| **Hypertension** | 1.47<br>(1.38-1.55) | 1.06<br>(1.02-1.10) | 1.35<br>(1.20-1.51) | 1.13<br>(1.07-1.20) |
| **Peripheral vascular disease** | 2.03<br>(1.75-2.35) | 1.34<br>(1.16-1.54) | 1.84<br>(1.34-2.52) | 1.48<br>(1.23-1.79) |
| **Stroke** | 1.79<br>(1.61-1.99) | 1.35<br>(1.23-1.48) | 1.77<br>(1.42-2.20) | 1.54<br>(1.36-1.74) |
| **Valvular disease** | 2.65<br>(2.31-3.03) | 0.90<br>(0.77-1.06) | 3.09<br>(2.38-4.00) | 1.41<br>(1.16-1.70) |
| **Venous thromboembolic disease** | 1.43<br>(1.28-1.60) | 1.23<br>(1.13-1.33) | 1.38<br>(1.11-1.72) | 1.26<br>(1.12-1.42) |

\*Fully adjusted models include age (modelled by use of restricted cubic splines with five knots for infection death, four knots for all other analyses), sex, socioeconomic deprivation, ethnicity, smoking, obesity, respiratory disease, diabetes, cancer, liver disease, kidney disease, neurological disease, psychiatric disease, rheumatological disease, abdominal aortic aneurysm, atrial fibrillation/flutter, coronary artery disease, heart failure, hypertension, peripheral vascular disease, stroke, valvular disease, venous thromboembolic disease. CV=cardiovascular.

**Table 4:** **Cancer deaths by sites**

|  | **Cancer death** |  |  |  |  |  |
| --- | --- | --- | --- | --- | --- | --- |
| **CV comorbidity** | **Breast** | **Colorectal** | **Lung & bronchus** | **Prostate** | **Uterine** | **Other** |
| No CV comorbidities | 820 (9.83%) | 823 (9.87%) | 1360 (16.31%) | 441 (5.29%) | 118 (1.42%) | 4776 (57.28%) |
| One CV comorbidity | 318 (6.32%) | 469 (9.32%) | 919 (18.27%) | 324 (6.44%) | 95 (1.89%) | 2906 (57.76%) |
| Two CV comorbidities | 55 (4.51%) | 90 (7.38%) | 273 (22.38%) | 83 (6.8%) | 12 (0.98%) | 707 (57.95%) |
| Three or more CV comorbidities | 4 (1.45%) | 28 (10.18%) | 69 (25.09%) | 18 (6.55%) | 3 (1.09%) | 153 (55.64%) |

CV=cardiovascular

**Table 5:**
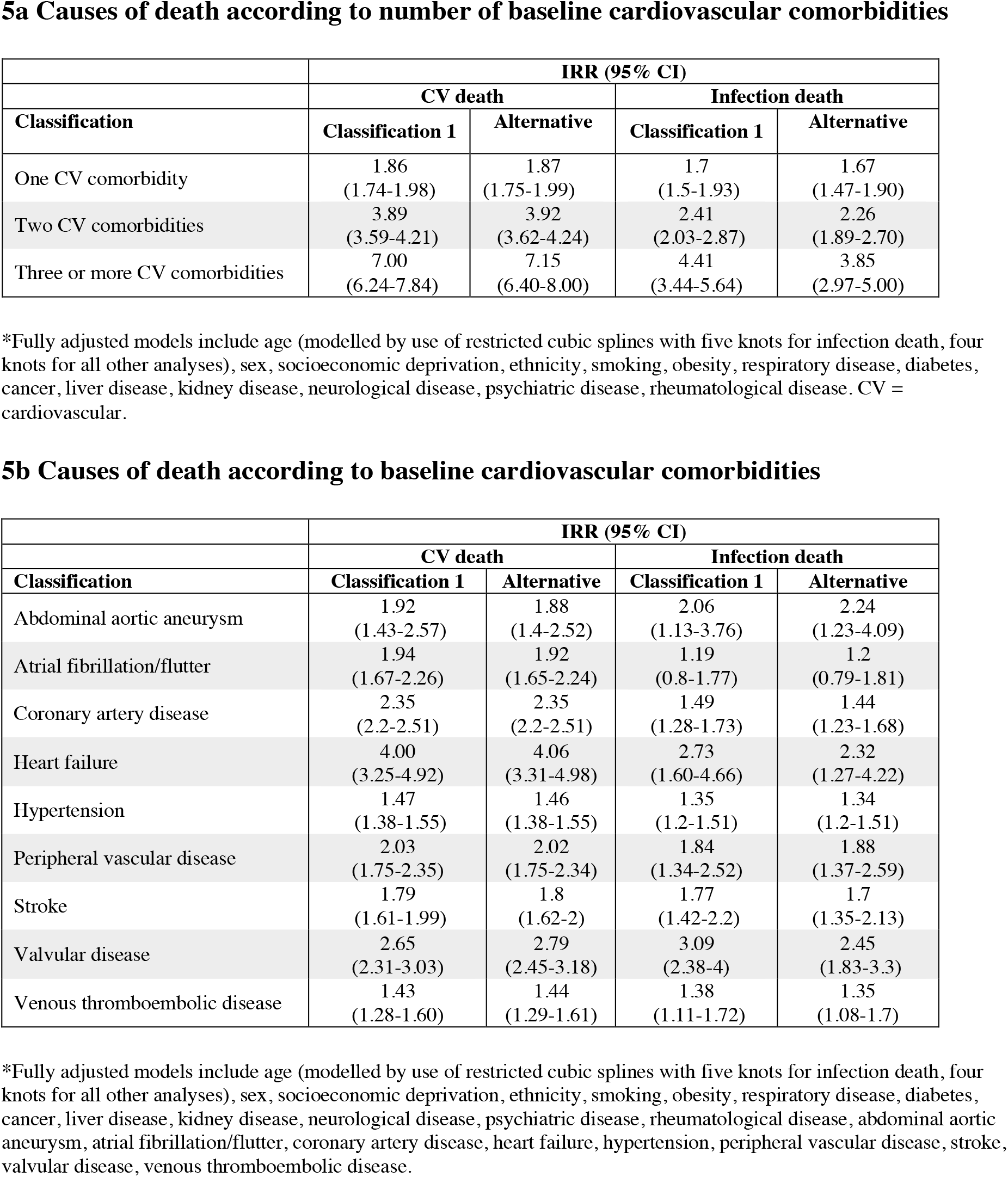
Cardiovascular and infection death classification sensitivity analyses. Total of 61 death events occurred that were reclassified for alternative classification.

#### Disease definitions

| **Disease** | **UK Biobank self-reported illnesses/operation included** |
| --- | --- |
| Abdominal aortic aneurysm | Aortic aneurysm<br>Aortic aneurysm rupture<br>Aortic aneurysm/repair or stent |
| Atrial fibrillation/flutter | Atrial fibrillation<br>Atrial flutter |
| Coronary artery disease | Angina<br>Coronary angioplasty (ptca) +/- stent<br>Coronary artery bypass grafts (cabg)<br>Heart attack/myocardial infarction<br>Triple heart bypass |
| Heart failure | Cardiomyopathy<br>Heart failure/pulmonary odema<br>Hypertrophic cardiomyopathy (hcm / hocm) |
| Hypertension | Essential hypertension<br>Hypertension |
| Peripheral vascular disease | Amputation of foot<br>Amputation of leg<br>Amputation of toe<br>Arterial embolism<br>Fem-pop bypass/leg artery bypass<br>Leg artery aneurysm repair<br>Leg artery angioplasty +/- stent<br>Leg claudication/ intermittent claudication<br>Other amputation<br>Peripheral vascular disease |
| Stroke | Brain haemorrhage<br>Ischaemic stroke<br>Stroke<br>Subarachnoid haemorrhage |
| Valvular disease | Aortic regurgitation / incompetence<br>Aortic stenosis<br>Aortic valve disease<br>Aortic valve repair/replacement<br>Heart valve problem/heart murmur<br>Heart valve surgery<br>Mitral regurgitation / incompetence<br>Mitral stenosis<br>Mitral valve disease<br>Mitral valve prolapse<br>Mitral valve repair/replacement<br>Other valve repair/replacement |
| Venous thromboembolic disease | Deep venous thrombosis (dvt)<br>Pulmonary embolism +/- dvt |

All comorbidities are defined using self-reported illness/operation at verbal nurse led interview (UK Biobank data fields 20002 and 20004).
Diseases also adjusted for in modelling include: obesity (defined using BMI) and self-reported: chronic respiratory disease, diabetes, chronic liver disease, chronic kidney disease, other neurological disease, psychiatric disorder, and chronic inflammatory and autoimmune rheumatological disease as defined previously<sup>2</sup>.

